# Explainable machine-learning model to classify culprit calcified carotid plaque in embolic stroke of undetermined source

**DOI:** 10.1101/2024.10.25.24316081

**Authors:** Yu Sakai, Jiehyun Kim, Huy Q Phi, Andrew C Hu, Pargol Balali, Konstanze V Guggenberger, John H Woo, Daniel Bos, Scott E Kasner, Brett L Cucchiara, Luca Saba, Zhi Huang, Daniel Haehn, Jae W Song

**Affiliations:** University of Pennsylvania; University of Massachusetts Boston; Drexel University; University Hospital of Würzburg; Hospital of the University of Pennsylvania; Erasmus MC; Universisty of Cagliari

## Abstract

**Background:** Embolic stroke of undetermined source (ESUS) may be associated with carotid artery plaques with <50% stenosis. Plaque vulnerability is multifactorial, possibly related to intraplaque hemorrhage (IPH), lipid-rich-necrotic-core (LRNC), perivascular adipose tissue (PVAT), and calcification morphology. Machine-learning (ML) approaches in plaque classification are increasingly popular but often limited in clinical interpretability by black-box nature. We apply an explainable ML approach, using noncalcified plaque components and calcification features with SHapley Additive exPlanations (SHAP) framework to classify calcified carotid plaques as culprit/non-culprit.

**Methods:** In this retrospective cross-sectional study, patients with unilateral anterior circulation ESUS who underwent neck CT angiography and had calcific carotid plaque were analyzed. Calcification-level features were derived from manual segmentations. Plaque-level features were assessed by a neuroradiologist blinded to stroke-side and by semi-automated software. Calcifications/plaques were classified as culprit if ipsilateral to stroke-side. Eight baseline ML models were compared. Three CatBoost models were trained: Plaque-level, Calcification-level, and Combined. SHAP was incorporated to explain model decisions.

**Results:** 70 patients yielded 116 calcific carotid plaques (60 ipsilateral to stroke; 270 calcifications (146 ipsilateral)). 17 plaque-level and 15 calcification-level features were extracted. Baseline CatBoost model outperformed other models. Combined model achieved test AUC 0.77 (95% CI: 0.59-0.92), accuracy 0.82 (95% CI: 0.71 - 0.91), mean cross-validation AUC 0.78. Plaque-level and calcification-level models performed lower (AUC 0.41 95% CI: 0.15-0.68, 0.60 95% CI 0.44-0.76). Combined model utilized five features: plaque thickness, IPH/LRNC volume ratio, PVAT volume, calcification minimum density, and total calcification volume over mean density ratio. Plaque thickness was most important feature based on SHAP values, with potential threshold at >2.6 mm.

**Conclusions:** ML model trained with noncalcified plaque and calcification features can classify culprit calcific carotid plaque with greater accuracy than models trained using only plaque-level or calcification-level features. Model using clinically interpretable features with SHAP framework provides explanations for its decisions and allows identification of potential thresholds for high-risk features.

Graphic Abstract
Overall design of our study.

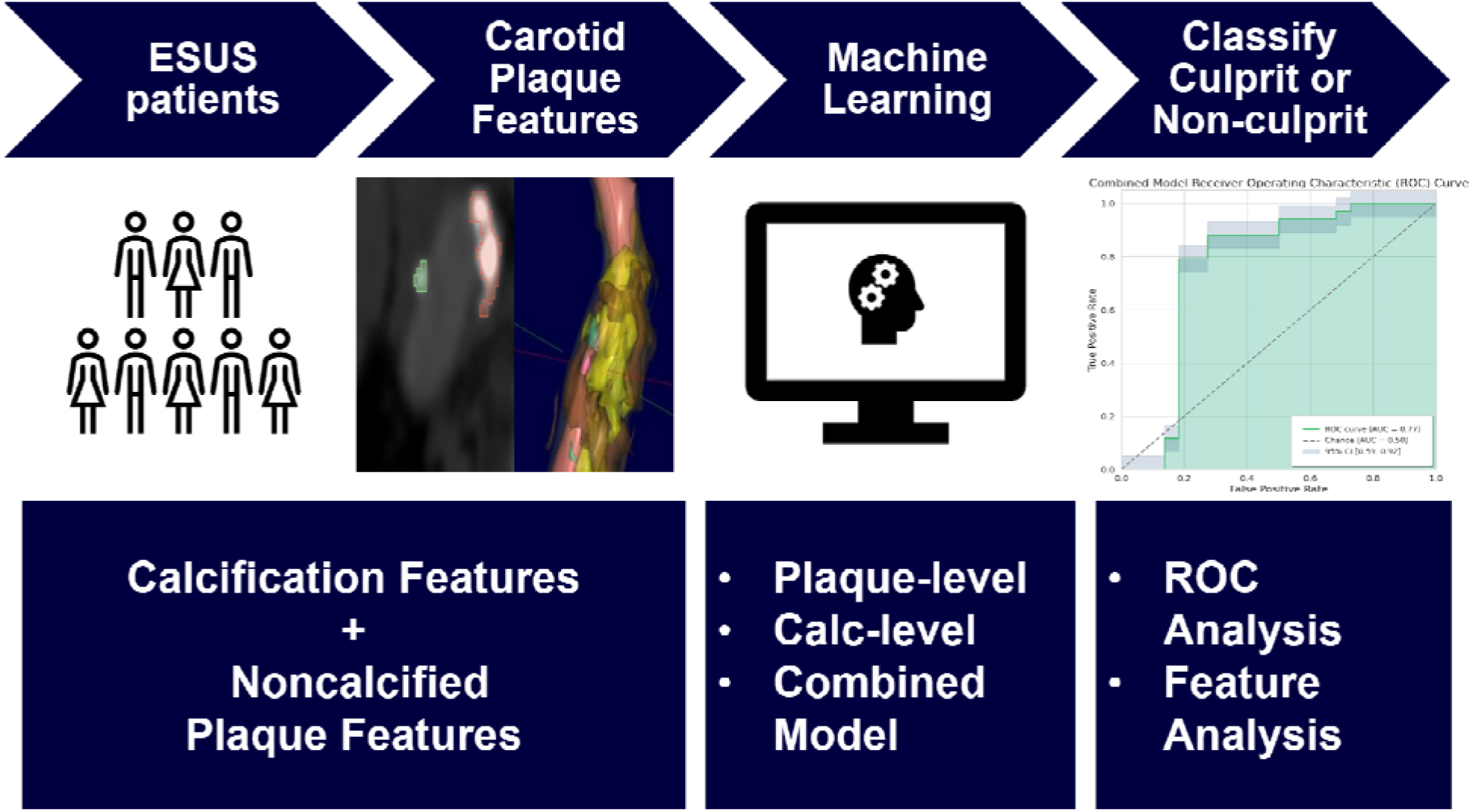

## Introduction

Embolic stroke of undetermined source (ESUS) presents significant diagnostic and therapeutic challenges due to its multifactorial etiology [1], affecting approximately 1 in 6 ischemic stroke patients, with a recurrence rate of 5% per year despite antithrombotic therapy [2].

Growing evidence suggests carotid plaques with <50% stenosis play a critical role in ESUS. The NAVIGATE-ESUS trial failed to demonstrate efficacy or safety of rivaroxaban compared with aspirin in patients with ESUS [3], potentially due to high enrollment (40%) of patients with carotid plaques <50% stenosis. The trial was stopped early and results did not reach statistical significance, but patients with carotid plaques showed a strong trend of higher stroke recurrence rates (5.4 vs. 4.3/100 patient-years) and a trend favoring aspirin over rivaroxaban (recurrent stroke rate 4.9 versus 5.9/100 patient-years) [4].

Studies suggest plaque vulnerability is complex and various imaging features should be considered for risk assessment [5]. For example, noncalcified intraplaque components, such as intraplaque hemorrhage (IPH) is a high-risk feature of carotid plaque associated with ischemia [6]. Meta-analysis demonstrated that recurrent stroke/TIA in patients with carotid plaques of <50% stenosis was 2.6/100 person-years and increased to 4.9/100 person-years if intraplaque hemorrhage was present [7]. Additionally, plaque calcification morphologies may also play a role in plaque stability, as suggested by imaging biomarkers such as “spotty calcification” and “rim-sign” [8, 9]. Furthermore, the presence of inflamed adipose tissue surrounding the carotid artery has also been identified as a possible indicator of plaque vulnerability [10]. These findings highlight the multifactorial nature of plaque vulnerability and consideration of a combination of these plaque composition and plaque calcification features may provide insight about high-risk plaque.

Supervised machine-learning (ML) models, a subfield of artificial intelligence, excel in addressing focused problems that involve high-dimensional data, especially binary classification tasks [11], and have been previously applied to stroke imaging [12]. The improved predictive accuracy of ML models often comes at the cost of increased complexity, resulting in “black box” approaches that obscure how decisions are made; this lack of transparency limits their adoption in healthcare, where explainability is crucial. Consequently, there is increasing demand for succinct, “explainable” models based on clinically interpretable features [13].

We hypothesize that an explainable ML model trained on a combination of clinically interpretable noncalcified plaque features and individual calcification features will be able to more accurately classify culprit and non-culprit calcific plaques in ESUS patients compared to a model incorporating only carotid plaque or plaque-calcification features.

## Materials and Methods

This study was deemed exempt by the University of Pennsylvania Institutional Review Board (853355). Individual patient consent was waived because of the retrospective nature of the study. The design and reporting of this study was completed according to the Strengthening the Reporting of Observational Studies in Epidemiology (STROBE) guidelines [14] and the Checklist for Artificial Intelligence in Medical Imaging (CLAIM) [15] (CLAIM checklist available as supplemental **Table S1**).

### Data Availability

The data and model can be made available on reasonable written request to the corresponding author. The ML model-building methodology described in this article is based on open-source resources, and the hyperparameters of our final models are shared in the Results.

### Patient Selection

This was a single-center retrospective cross-sectional study. Patients satisfying the following criteria were included: consecutive patients with unilateral anterior circulation acute ischemic stroke admitted to our health system between October 1, 2015 – April 1, 2017, of ≥18 years of age, who underwent CT angiography (CTA) of the neck within 10 days of stroke onset, met criteria for ESUS as determined by vascular neurologists, and confirmation of carotid plaque calcifications on CTA determined by visual inspection. ESUS was confirmed following method prior reported [16]. The following patients were excluded: patients with multiple possible mechanisms of ischemic stroke, evidence of simultaneous acute ischemic stroke in more than one vascular territory, prior carotid endarterectomy or stenting, occlusion of either cervical internal carotid artery.

### Definition of Culprit vs. Non-culprit plaque/calcification

Cervical carotid plaques and associated calcifications were defined as culprit or non-culprit if they were ipsilateral or contralateral to the stroke-side, respectively.

### Imaging Acquisition

CTA of the neck was acquired in the axial plane using a 4th generation, helical CT scanner, slice thickness ranging from 0.625 to 1.5 mm. Iodinated contrast (100mL Isovue-370) was administered intravenously through a 20-gauge (or larger) antecubital catheter.

### Image Analysis

All image segmentations and scoring were performed blinded to stroke-side. To obtain plaque-level features, a board-certified neuroradiologist (JW) reviewed the neck CTAs and assessed for carotid plaque thickness, presence of ulcerations, and degree of stenosis based on NASCET criteria. Cases with greater than 50% stenosis ipsilateral to stroke were excluded; sub-analysis showed contralateral plaques also had less than 50% stenosis (median 0%, IQR 0-15%). A semi-automated plaque composition segmentation software (Elucid Bioimaging, Boston, MA) was used to quantify intraplaque hemorrhage (IPH), lipid rich necrotic core (LRNC), plaque matrix and perivascular adipose tissue (PVAT) volumes [17] and checked by a neuroradiologist (JWS). For plaque-calcification level features, CTA necks were screened visually for calcific carotid bifurcation plaques across a 4 cm segment of the carotid bifurcation by two independent evaluators (ACH, HQP) with consensus by radiologists (YS, JWS); cases without carotid plaque calcifications by visual inspection were excluded. Each carotid artery was evaluated independently. If a patient had only one carotid artery with calcified plaque, then that plaque was used for further segmentation of the calcification and the carotid artery without calcified plaque was excluded. Two evaluators (ACH, HQP) independently labeled each discrete plaque calcification using 3D Slicer (https://www.slicer.org) and both labelling masks were overlaid and proofed by a radiologist (YS) to curate the final labeled dataset. Segmentation maps from 3D Slicer were used to derive calcification morphological features: volume, diameter, surface area, roundness, elongation, flatness, and density (attenuation values in Hounsfield units (HU)). Each discrete plaque calcification was manually scored by an evaluator (PB) for the location of the calcification relative to the vessel wall (adventitial, intimal, or transmural) and angle of calcification arc (between 0-90°, 90-180°, 180-270°, or 270-360°). “Spotty calcification” was defined as calcifications with arc <90° and axial thickness <3mm [8, 18]. “Rim-sign” was identified as a plaque-level feature designated by the presence of a calcification at an adventitial site with >90 degrees arc with adjacent non-calcified plaque measuring ≥2mm in thickness [9].

### Statistical Analysis and Machine Learning (ML) Methods

All statistical analyses and ML methods were conducted with Python (3.10.12) using relevant packages: scipy 1.13.1; numpy 1.26.4; matplotlib 3.7.1; pandas 2.1.4; scikit-learn 1.3.2; xgboost 2.1.1; catboost 1.2.7; lightgbm 4.5.0).

#### Input Data for ML models

Three datasets were prepared: (1) Plaque-level: features and laterality of each plaque relative to stroke; (2) Calcification-level: features and laterality of each calcification; (3) Combined: Merged Plaque-level and Calcification-level features, each row representing a calcification and its parent plaque. Baseline feature distributions were assessed using the Wilcoxon rank sum or t-test with normality evaluated by the Shapiro-Wilk test.

Training and Test Set Split: To simulate a prospective use case, data was split by admission date: cases from 2015-2016 formed the Training set and cases from 2017 formed the Test set.

#### Feature Engineering and Selection

Feature engineering and selection were performed on the Training set. Continuous variables were discretized into ten quantiles to reduce noise [19]; thresholds based on Training set were applied to the Test set preventing data leakage [20]. Chi-square tests evaluated associations between binned features and a binary outcome (ipsilateral vs. contralateral). Monte Carlo simulations (10,000 iterations) ensured reliable p-values when expected counts were <5. Plaque-level and Calcification-level models used significant features (p<0.05) if available. Five clinically relevant features were selected for the Combined model (see Results).

#### Baseline ML Algorithm Type Assessment and Selection

Eight common ML algorithms [21–24] —Logistic Regression, SVM, Decision Tree, Random Forest, Naïve Bayes, XGBoost, CatBoost, and LightGBM—at default settings were compared using the Combined model Training set. CatBoost, showing highest performance through 10-fold cross-validation, was selected for further optimization.

#### Training

Plaque-level, Calcification-level, and Combined CatBoost models [22] were trained. *Cross-validation and Hyperparameter tuning:* CatBoost hyperparameters were optimized using exhaustive grid search (GridSearchCV) with 10 group-fold cross-validation, minimizing logarithmic loss. Three hyperparameters were tuned: depth (3-5), iterations (100-300), and learning rate (0.001, 0.01, 0.1); descriptions of hyperparameters have been previously described [22]. Group-fold cross-validation [25] based on Plaque ID ensured no calcifications from the same plaque appeared in both training and validation sets, preventing data leakage. Mean cross-validation AUC with 95% confidence interval was reported.

#### Testing and Performance Analysis

Final models were evaluated on their respective Test datasets. Performance metrics included accuracy, ROC curve, and AUC. 95% confidence intervals were computed using the bootstrap method (1000 iterations) [26]. AUCs between models tested on separate test sets were compared using the independent t test. Additional metrics such as precision, recall, and F1-score were also calculated for the best model.

Feature Analysis:

### Feature correlation analysis

Spearman’s rank correlation matrix was generated for the Combined model features.

### SHAP analysis

SHapley Additive exPlanations values (SHAP) were calculated for each feature for the final best model [27]. SHAP values help visualize the contribution of each feature to the model’s predictions and explain directionality thereby making “black-box” ML models interpretable [28, 29].

#### Failure Analysis

A confusion matrix for the best model’s performance on the test set was generated. SHAP waterfall plots were used to visualize how specific features contributed to errors in misclassified cases.

## Results

### Patient demographics

Patient flow diagram is shown in **Figure 1** and demographic data are summarized in **Table 1**. From 772 acute ischemic stroke patients, 94 met criteria for ESUS among which 24 were excluded due to the absence of carotid plaque calcifications, as a study aim was to investigate and compare a model with plaque calcification-level features. 70 patients (50% men, mean age 68+/-11 yeas) were included in this study.

**Figure 1.**
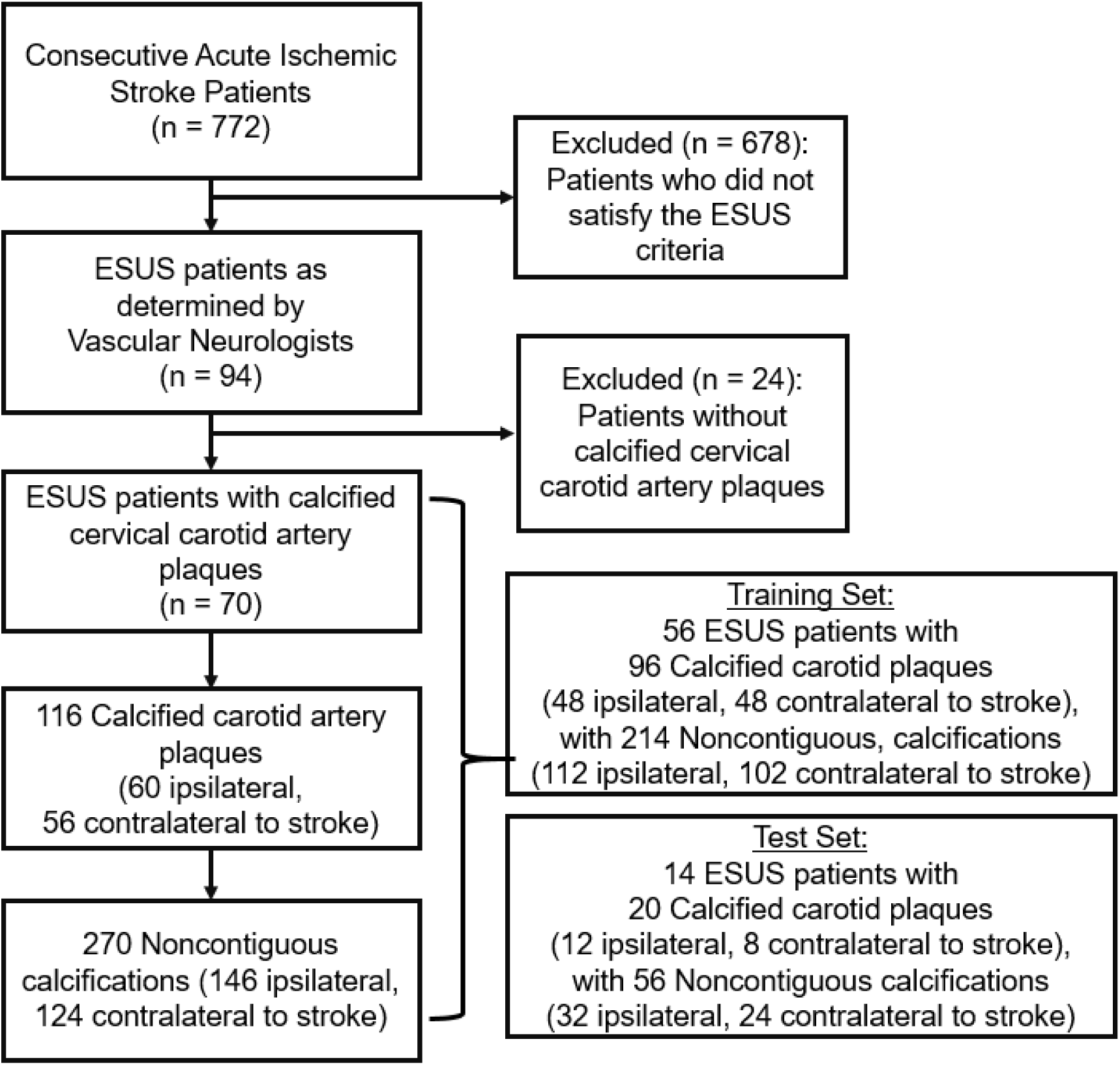
Patient flow diagram with Train and Test split. Flow diagram shows 70 total embolic stroke of undetermined source (ESUS) patients included in this study which yielded 116 calcified carotid artery plaques and 270 noncontiguous calcifications. This was split into Training (56 patients with 86 calcified carotid artery plaques and 214 noncontiguous calcifications) and Test sets (14 patients with 20 calcified carotid plaques and 56 noncontiguous calcifications).

**Table 1.** Demographic data.

| Total Dataset<br>N= 70 |  |  | Training Set<br>N=56 | Test Set<br>N=14 |
| --- | --- | --- | --- | --- |
| Characteristic | Mean<br>(SD) | n (%) |  |  |
| Age | 68 (11) |  | 67 (10) | 71 (14) |
| Sex |  |  |  |  |
| Female |  | 35 (50) | 26 (46%) | 9 (64%) |
| Diabetes |  | 22 (31%) | 18 (32%) | 4 (29%) |
| Hypertension |  | 53 (76%) | 44 (79%) | 9 (64%) |
| Coronary Artery Disease |  | 19 (27%) | 16 (29%) | 3 (21%) |
| Dyslipidemia |  | 28 (40%) | 18 (32%) | 10 (71%) |
| Smoker |  | 14 (20%) | 12 (21%) | 2 (14%) |

Abbreviations: SD: standard deviation.

### Imaging Feature Derivation

From 70 ESUS patients, 116 calcific carotid plaques were identified on neck CTA (60 ipsilateral, 56 contralateral to stroke). Each carotid artery was evaluated independently. 270 noncontiguous plaque calcifications were manually segmented (146 ipsilateral, 124 contralateral) (**Figure 1**). 17 plaque-level and 15 calcification-level features were derived, definitions summarized in supplemental **Table S2.**

Shapiro-Wilks test showed features were not in normal distribution (p<0.05) except for minimum calcification density (p = 0.36). The distributions of the imaging features for the Combined dataset (N=270) between ipsilateral (N=146) and contralateral (N=124) sides are summarized in **Table S3.** During the model-building process and feature selection, distribution information of the entire dataset (which includes information about the test set) were not leveraged.

### Training and Test Split

The Training set (2015-2016 admissions) yielded 56 patients with 96 calcified carotid artery plaques (48 ipsilateral) with 214 noncontiguous calcifications (112 ipsilateral). The Test set (2017 admissions) yielded 14 patients with 20 calcified carotid artery plaques (12 ipsilateral) with 56 noncontiguous calcifications (32 ipsilateral) (**Figure 1**). This naturally provided a ∼80:20 split. Demographic data of Training/Test sets are provided in **Table 1**.

### Feature Engineering and Selection

Significant features based on Chi-square analysis with Monte Carlo simulation after 10-quartile binning are summarized in supplemental **Table S4**.

Twelve significant plaque-level features were used to train the Plaque-level model (supplemental **Table S4**). There was no significant calcification-level feature and thus all calcification features were used for the Calcification-level model (Calc-level features in supplemental **Table S2**). Minimum Density approached significance (p=0.056) and was thus included for consideration for the Combined model. Pearson correlation matrix of these 13 features showed high correlations between many features (supplemental **Figure S1**). From 13 features, 5 were selected for the Combined model based on domain knowledge and multicollinearity analysis (see **Supplement: Feature Selection**): IPH over LRNC volume ratio (IPH_vol_/LRNC_vol_), Plaque thickness (mm), Perivascular adipose tissue (PVAT) volume (mm^3^), Plaque-level total calcification volume over mean density (mm^3^/HU), and Calcification minimum density (HU). Spearman’s rank correlation matrix showed that there were no strong correlations among these five features (**Figure 2**).

**Figure 2.**
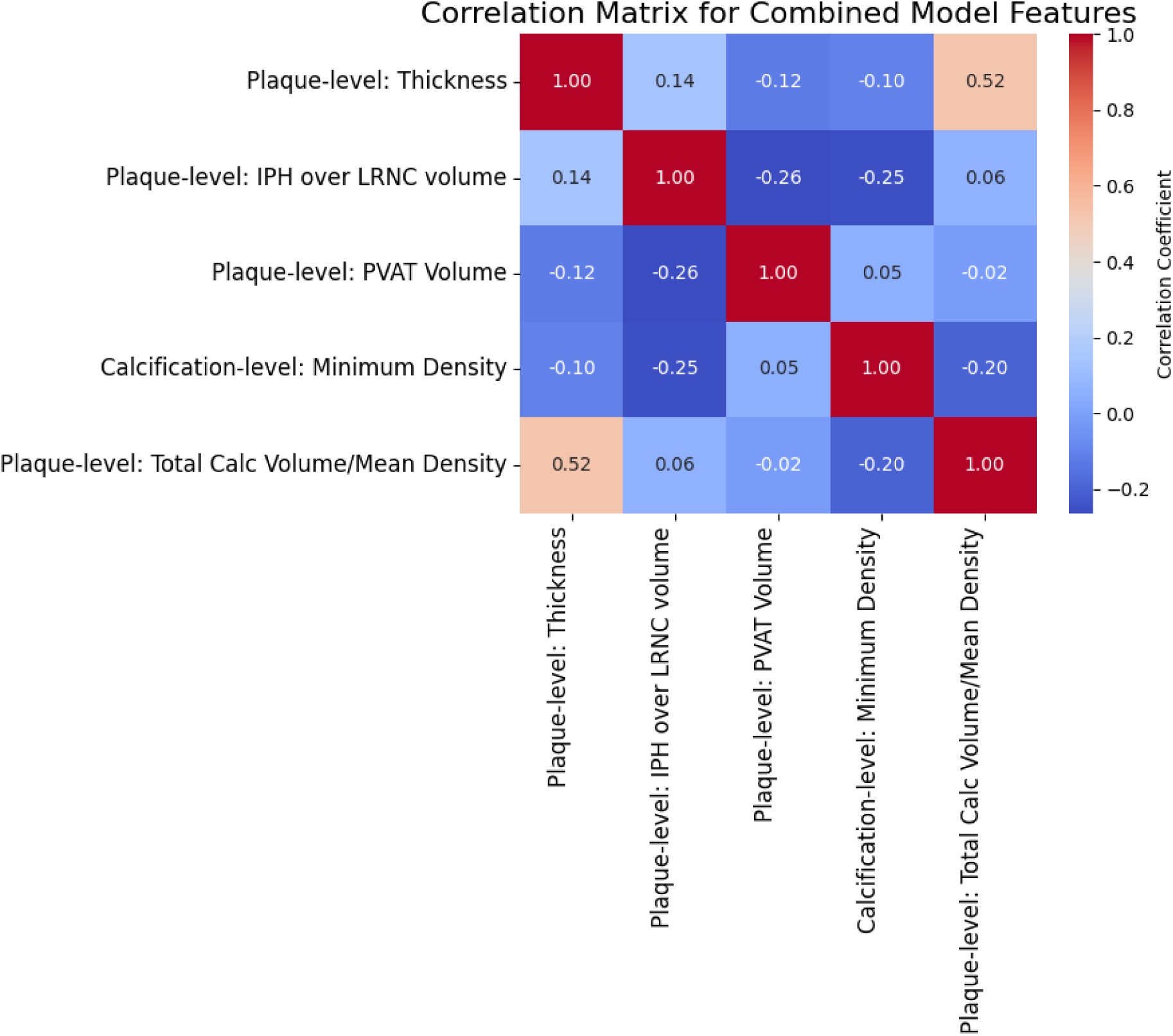
Spearman’s rank correlation matrix for Combined model features. Spearman’s rank correlation matrix showed that for the five features used by the Combined model, there were no features with strong correlation in the Training Set (correlation coefficient >0.7). Abbreviations: IPH: intraplaque hemorrhage, LRNC: lipid-rich-necrotic-core, PVAT: perivascular adipose tissue.

### Baseline model performance

Comparisons of eight commonly employed models in their default parameter settings using the Combined dataset demonstrated that all model types struggled to perform greater than chance, but CatBoost showed highest performance with AUC of 0.62 (**Figure 3**).

**Figure 3:**
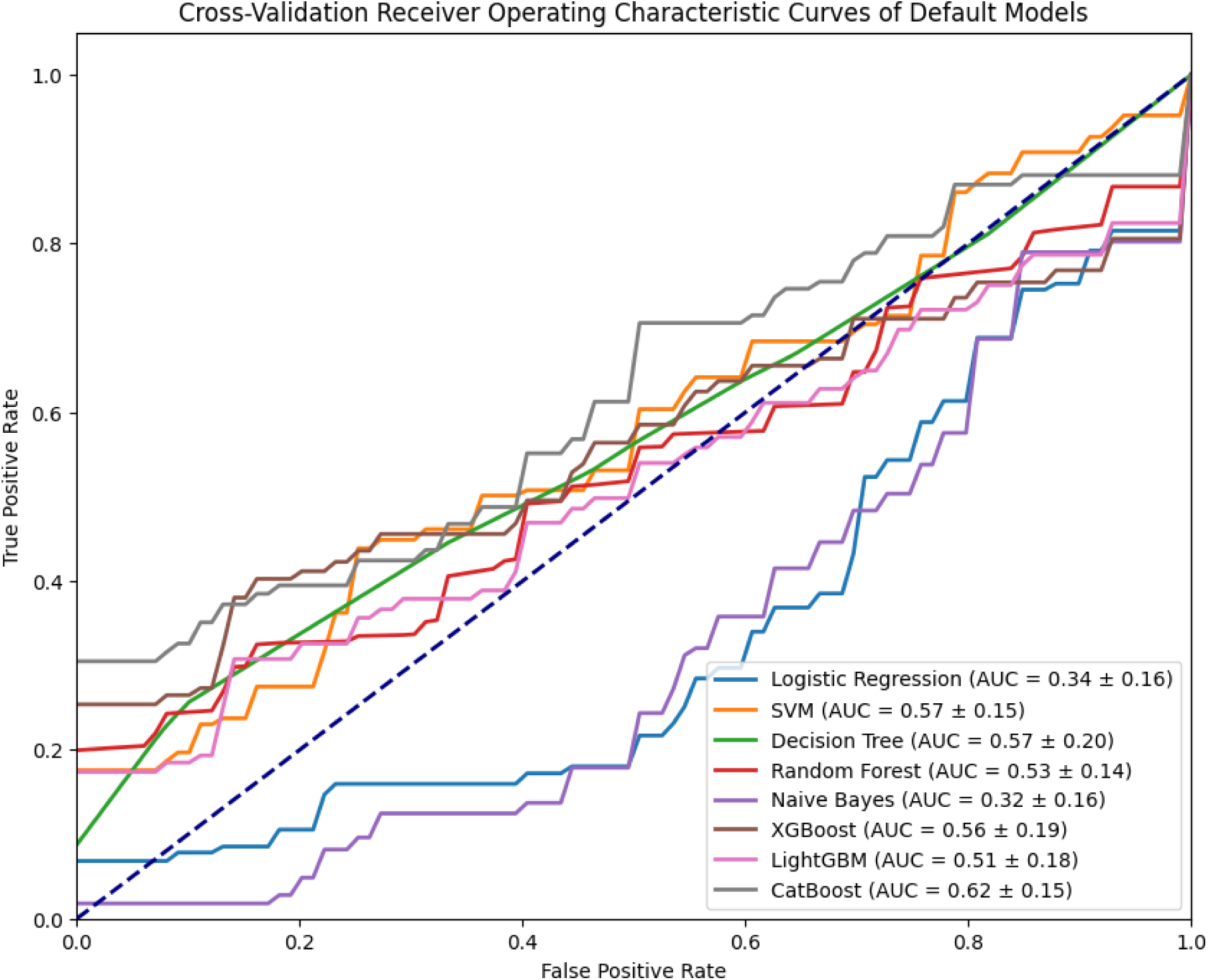
Receiver operating characteristic curves of eight default ML models. Receiver operating characteristic curves obtained during cross-validation process (thus using only Training set, not Test set) shows that eight models in their default settings struggled to classify culprit and non-culprit plaques better than chance, but CatBoost (gray) performed highest (AUC 0.62). Abbreviations: AUC: Area under the curve, CatBoost: categorical boosting, SVM: support vector machine, LightGBM: light gradient boosting machine, ML: machine learning, XGBoost: extreme gradient boosting.

### Hyperparameter-tuning of models

The optimal hyperparameter values for the Combined model were learning_rate 0.001, depth 5, and iterations 200. Hyperparameters for the Plaque-level and Calcification-level models are in supplemental **Table S6**.

### Final model performance

The Combined model achieved mean cross-validation AUC of 0.78, test set accuracy 0.82 (95% CI: 0.71-0.91) and AUC 0.77 (95% CI 0.59-0.92) (**Figure 4**).

**Figure 4.**
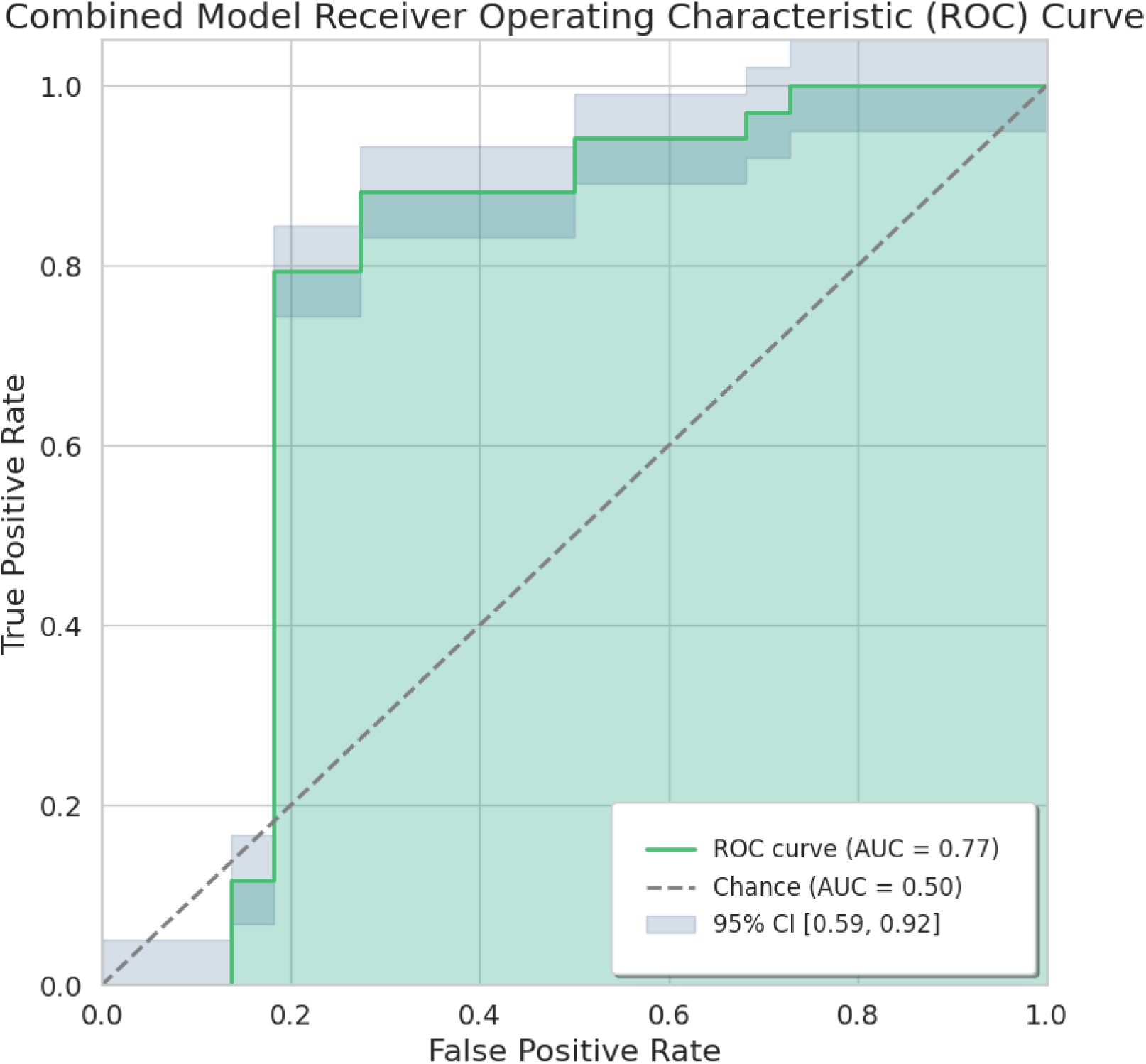
Receiver operating characteristic curve of Combined Model. Receiver operating characteristic curve of Combined model on test set shows AUC of 0.77 (95% CI 0.59-0.92). Abbreviations: AUC: Area under the curve. ROC: Receiver operating characteristic. CI: confidence interval.

Plaque-level and Calcification-level models performed lower (p<0.001), with AUC 0.41 (95% CI 0.15-0.68) and AUC 0.60 (95% CI 0.44-0.76), respectively. Additional metrics are summarized in supplemental **Table S7**, and Precision-Recall curve with AUC of 0.82 is in supplemental **Figure S2.**

Sub-analysis of aggregating the Combined model results from the calcification-and-plaque-pair-level results to the plaque-level resulted in AUC of 0.74, again outperforming the Plaque-level model.

### SHAP Feature Analysis of Best Performing Model (Combined Model)

Based on the mean SHAP score (represents impact of each feature in influencing the model to decide culprit or non-culprit), plaque thickness was the most important feature of the Combined model, followed by PVAT volume, total calcification volume to mean density ratio, IPH_vol_/LRNC_vol_ ratio, and calcification minimum density (**Figure 5**). Example of each imaging feature is shown in **Figure 6**.

**Figure 5.**
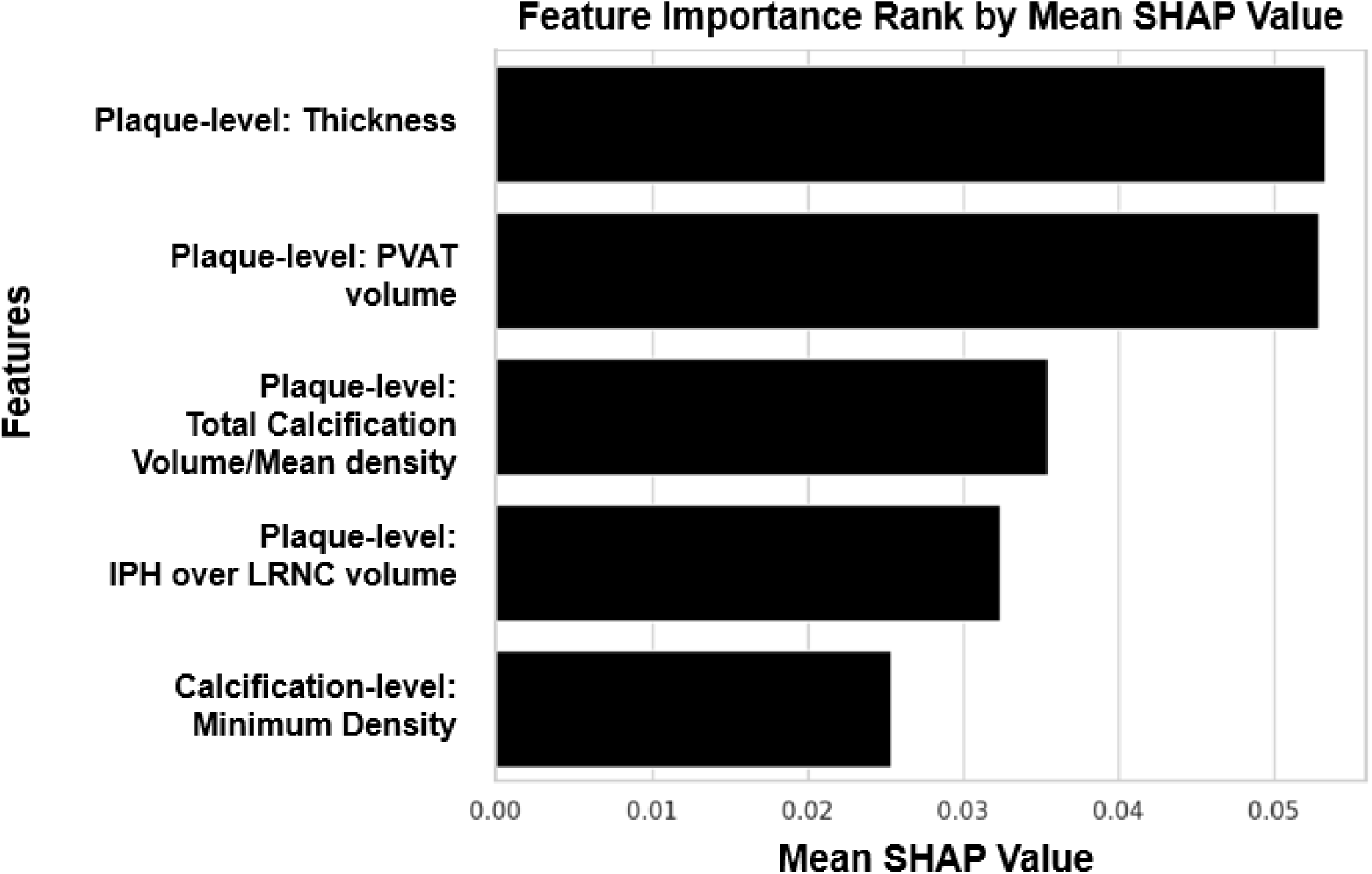
Feature importance ranking of Combined model. Five features used for the Combined model were ranked by mean SHapley Additive exPlanations (SHAP) score, with plaque thickness being the most important. Abbreviations: PVAT: Perivascular adipose tissue, IPH: Intraplaque hemorrhage, LRNC: Lipid-rich-necrotic-core.

**Figure 6.**
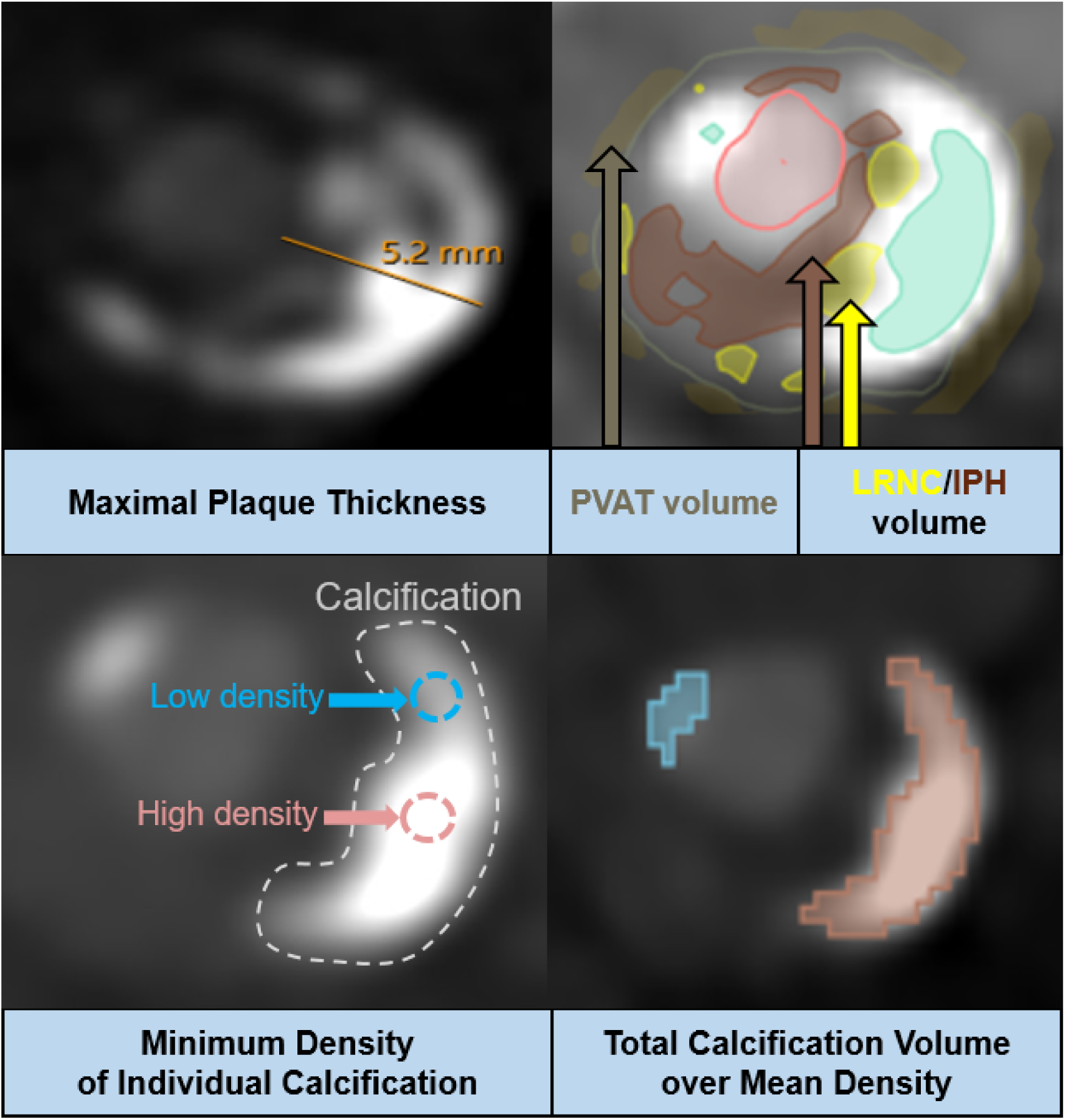
Imaging features used in the Combined model. Example of each imaging feature used in the Combined model; Maximal plaque thickness was measured from CTA image directly, Lipid-rich-necrotic-core (LRNC), intraplaque hemorrhage (IPH) and perivascular adipose tissue (PVAT) volumes are based on Elucid Bioimaging software, and calcification-related features are based on 3D Slicer segmentations.

SHAP dependency plots visualized how the model used each feature to determine culprit/non-culprit and allowed visualization of potential thresholds for the features (supplemental **Figure S7**) based on our 10 quantile bins (supplemental **Table S5**): Plaque thickness >2.6 mm, PVAT volume >112 mm^3^, total calcification volume to mean density ratio >0.32, and IPH_vol_/LRNC_vol_ ratio >0.4, and calcification minimum density >240HU.

Top 10 important features of the Calcification-level and Plaque-level models by mean SHAP scores are summarized in supplemental **Figures S3** and **S4**.

### Failure Analysis

The confusion matrix for the Combined model (supplemental **Figure S5**) showed 16 true negatives, 4 false negatives, 6 false positives, and 30 true positives. SHAP water-fall plots analyzed incorrect cases, and select examples are shown in supplemental **Figure S8**. In a false positive example (supplemental **Figure S8A**), the model incorrectly classified the calcification-plaque pair as culprit because there was relatively high plaque thickness and PVAT, overriding the impact by minimum calcification density. Conversely, in a false negative example (supplemental **Figure S8B**), the model incorrectly classified a case as non-culprit because there was relatively low plaque thickness and PVAT volume.

## Discussion

### ML approach to calcified carotid plaque classification

Our Combined model achieved an AUC of 0.77 (95% CI: 0.59-0.92) and a precision-recall AUC of 0.74 (95% CI: 0.58-0.90) for classifying culprit (ipsilateral) versus non-culprit (contralateral) calcification-plaque pairs in patients with <50% carotid stenosis. Comparable studies to date have focused on plaques >50% stenosis. For example, a ML model using clinical metrics, plaque stenosis and size classified culprit plaque with AUC of 0.73 [30], and another model trained on CT radiomics in a cohort with mean ipsilateral stenosis of 72% achieved AUC of 0.76 [31]. Two additional models using radiomic features reported AUCs >0.90 [32, 33], but these were in carotid endarterectomy candidates (>70% stenosis). Another recent study involving patients with >50% stenosis used CTA-based plaque compositions for a model with AUC of 0.89 [34]. In contrast, our study focusing on plaques with <50% stenosis may cause ischemic stroke via a different mechanism (e.g., plaque rupture and embolism) and leverages both noncalcified component and calcification features. Comparing eight different models at default-settings suggested that our dataset presented a challenging binary classification task (**Figure 3**). However, fine-tuning of the CatBoost model resulted in improved performance (**Figure 4**), consistent with prior research showing CatBoost may outperform other ML models, including in stroke-related research [35, 36]. Our Combined model also outperformed non-ML classification methods based on domain knowledge, such as plaque-thickness (>3 mm; AUC 0.63), presence of IPH (AUC 0.53), “spotty calcifications” (AUC 0.50), and “rim-sign” (AUC 0.50), and combination of these criteria (AUC 0.46), underscoring the advantages of a ML approach (details in **Supplement)**.

### Explainable ML model with 5 clinically-relevant features

There is growing demand for explainable ML models over high-performing “black-box” models, particularly in healthcare, where trust and subsequent adoption is closely tied to model interpretability [13]. Models trained on radiomic features—quantitative metrics from medical images—can achieve high performance but are difficult to interpret due to complex features such as “entropy” [32] or “Grey Level Dependence Matrix: dependence Variance” [31]. Our model provides five explainable features with SHAP analysis to clarify the decision-making process and identify potential thresholds for each feature.

### Plaque Thickness

A study comparing ESUS and cardioembolic stroke cohorts found plaque thickness was significantly greater ipsilateral to stroke than contralateral in ESUS but not in cardioembolic strokes [37]. While there is no established cutoff, studies have reported that plaques with <50% stenosis with >3 mm thickness are more prevalent on ipsilateral side in ESUS patients [16, 38]. Our model and cutoff of >2.6 mm aligns with these findings.

### IPH/LRNC volume

IPH is a key marker of carotid plaque vulnerability with strong association with stroke risk as highlighted by two recent meta-analyses [39, 40]. LRNC, composed of cholesterol crystals and necrotic cell debris, is also associated with stroke [40, 41], and the ratio of IPH/LRNC volume has further been tied to stroke risk [42]. In a prior ML model [25], this ratio was the most important feature, with top five features also including IPH volume/percentage and lipid minus IPH volume/percentage. For our Combined model, we incorporated the ratio of IPH/LRNC volume. Our model suggested a potential cutoff at 0.40 for identifying culprit cases, aligning with the prior finding of 0.50 [25].

### PVAT volume

PVAT, the adipose tissue surrounding vessels, is involved in regulating vascular homeostasis [43]. There is growing evidence that PVAT may relate to plaque inflammation [10, 44], which makes it a potential imaging biomarker in ESUS [45]. Prior models using plaque compositions did not include PVAT [34]. Our model suggests higher PVAT volume relates to culprit plaque (supplemental **Figure S6**). Our model suggests 112 mm^3^ may be a threshold for risk stratification in ESUS. Further research on an external dataset is warranted.

### Features related to Calcification

The role of plaque calcification density on stability and its relationship with calcification volume remain areas of investigation [46]. Studies in coronary arteries showed that when considering calcium density alone, higher density is associated with increased cardiovascular risk [47]. Our model appears to align with this, as cases with higher minimum calcification density were more often classified as culprit. However, other coronary artery studies showed that after adjusting for calcium area or volume, higher calcium density may be associated with lower risk of events [48, 49]. Given our model classified higher minimum density and a higher ratio of calcification volume to density as culprit, the effect of increased calcification volume may be stronger in our dataset. Further studies are warranted to explicitly study the interaction between calcification density and volume with respect to plaque stability. In our cohort, previously reported high-risk features of “spotty calcifications” and the plaque-level “rim-sign” were not successful in accurately classifying culprit calcification-plaque pairs (see **Sub-analysis of Non-ML methods** in **Supplement**).

### Limitations

There are several limitations. First, our ML models were trained with and tested on a small-sample (N=70 patients) single-center data. While our data and model output generally agreed with the literature, ulcerations in our data were rarely observed and present on the carotid plaque contralateral to stroke-side. Ulceration is a well-established high-risk carotid plaque feature and is expected to be more prevalent ipsilateral to stroke-side. This is likely due to variability in small sample size. Ulceration was removed during the feature selection process and was also not included in recent similar work [34]. For future work in an independent validation cohort, it may be reconsidered as an additional potential model feature.

Second, carotid plaque calcifications were manually segmented by visual assessment, which may have introduced bias. However, we had 3 independent evaluators, including expert proofing, for this time-intensive task to create the segmentation maps. Additionally, a fourth evaluator also reviewed all maps and scored each calcification to determine arc angle and location. Nonetheless, at the voxel-level there may have been over/under-segmentations. Effects due to data noise and outlier values were mitigated by our quantile binning process.

Third, while we have explored eight different ML models (**Figure 3**), we did not compare our results to a convolutional neural network approach in which the segmentation maps and the underlying imaging data would be used as direct inputs. In a carotid endarterectomy population, a convolutional neural network model classified plaque using CT images with histological specimens as a ground truth and identified unstable plaque according to the modified American Heart Association definition with AUC of 0.97 [33]. While promising, the model was not validated in a stroke population with carotid plaque of <50% stenosis. While such a computer-vision approach may be a valid alternative, ML methods such as CatBoost may be preferred on a tabulated dataset because the feature importances are readily interpretable, especially when combined with the SHAP framework.

Our Combined model was more sensitive than specific in classifying culprit calcification/plaque features, resulting in more false positives than false negatives on the test set (supplemental **Figure S5**). Although our model achieved an AUC of 0.77, plaque thickness was the most important feature, which may lead to missed cases with low thickness but a high IPH-to-LRNC ratio. To avoid overfitting, external validation of the model is essential for improving sensitivity.

In this study, we screened ESUS cases for those with calcified carotid plaques of less than 50% stenosis. We assumed that this cohort experienced strokes due to ipsilateral plaques, but it is important to acknowledge that other stroke etiologies remain possible. While this assumption is a limitation of this model, literature suggests that it is less common for other etiologies to overlap in patients with carotid plaques [50].

### Clinical Implications

Using this unique ESUS dataset incorporating detailed plaque-level and calcification-level imaging features, we employed advanced ML techniques to classify calcified carotid plaques as either culprit or non-culprit using clinically-explainable CT imaging features. The five features used to build our Combined model parallel our understanding from imaging and histologic studies and include plaque thickness, IPH_vol_/LRNC_vol_, PVAT, minimum calcification density, and calcification volume/density ratio. Further investigations on these features may improve risk stratification in ESUS patients with carotid plaque with <50% stenosis. A longitudinal follow-up of ESUS patients with carotid plaques and calcifications classified as culprit by this model is a future direction to assess whether these features are truly predictive of future ischemic events.

## Conclusions

Our ML model trained on a combination of plaque-level and calcification-level metrics can classify culprit calcification-plaque pairs with AUC 0.77, better than models only trained with plaque-level or calcification-level features. We provide a succinct model with 5 unique clinically-relevant features with an explainable SHAP framework to interpret its decision-making process.

## Nonstandard Abbreviations and Acronyms

AUC: Area Under the Curve
CatBoost: Categorical Boosting
CTA: Computed Tomography Angiography
ESUS: Embolic Stroke of Undetermined Source
IPH: Intraplaque Hemorrhage
Light GBM: Light Gradient Boosting Machine
LRNC: Lipid-rich Necrotic Core
MATX: Plaque Matrix
ML: Machine Learning
PVAT: Perivascular Adipose Tissue
ROC: Receiver Operating Characteristics curve
SHAP: SHapley Additive exPlanations
SVM: Support Vector Machine
XGBoost: eXtreme Gradient Boosting

